# SARS-CoV-2 vaccines breakthrough infection hospitalizations after one dose in Libya: cohort study

**DOI:** 10.1101/2021.09.26.21263691

**Authors:** Mohamed Hadi Mohamed Abdelhamid, Iman A. Almsellati, Badereddin B Annajar, Alaa .H Abdelhamid, Hafsa .A Alemam, Mohammed Etikar

## Abstract

SARS-CoV-2 infection is widely spread over people, from youth to the elderly. Vaccination against SARS-CoV-2 is an important preventive measurement to help end the SARS-CoV-2 pandemic. From 30 April to 15 July, we collected the number of people infected with SARS-COV-2 and the mortality rate from daily reports issued by the National Center for Disease Control of Libya (NCDC). Approximately 445000 doses have been administered in Libya since 10 April, and 5 thousand doses are now being administered during this period on a daily basis. To estimate the rate of breakthrough vaccine infection of the SARS-COV-2 in Libya. We found that one dose of the three different types of vaccines had decreased the virus transmission across people and mortality rate until 10 weeks after the first dose. This study highlights the dramatic success of the early months of the nation’s coronavirus vaccines rollout.

## Introduction

SARS-CoV-2 infection is widely spread over people, from youth to the elderly. Mass vaccination operations to prevent coronavirus disease 2019 (Covid-19) are ensuing in 207 countries; 32% of the world population has received at least one dose of a SARS-CoV-2 vaccine, and 24% is fully vaccinated [1]. Approximately 4,84 billion doses have been administered worldwide, and 34,86 million are now administered daily [1,2]. Interestingly, SARS-CoV-2 several studies have found that most hospitalizations and deaths due to SARS-CoV-2 infection were among immunocompromised individuals, persons with co-morbidities, elderly and / or unvaccinated people [3–5].

Furthermore, previous studies showed that the vaccines reduce the risk of SARS-CoV-2 infection, especially illness severity, among partially vaccinated people [6–9] (Table 1).

**Table 1.** The efficacy of the vaccines against symptomatic, severe, and hospitalization SARS-CoV-2 infection.

| **Vaccines** | *SARS-CoV-2 infection* | *vaccines against symptoms*<br>*SARS-CoV-2 after receiving*<br>*the first dose* | *Against*<br>*hospitalization*<br>*after receiving*<br>*the first dose* |
| --- | --- | --- | --- |
| **AstraZeneca** | 67 % | 68.7% | 76% |
| **Sputnik-V** | 80% | 81% | 87% |
| **CoronaVac** | 49.6% | 70% | 99.2% |

Libya’s National Center for Disease Control (NCDC) reported that the first case of SARS-CoV-2 was identified in Libya on 24 Mars 2020 [10–12]. In addition, as reported by the World Health Organization (WHO), many countries are not very well-prepared to fight the pandemic such as (Libya, Iraq, and Yemen) these countries are more vulnerable to the impact of this pandemic [13].

Nevertheless, In Libya, the national immunization program launched a long-delayed SARS-CoV-2 vaccination program on 10 April 2021, through 400 immunization centers distributor in all cities of the country [12].

Currently, most cases of SARS-CoV-2 effects people who have not been fully vaccinated. Over the world, many hospitals have recorded cases of SARS-CoV-2 infection in patients after being fully vaccinated, this situation is called a vaccine breakthrough cases [2,14]. Although SARS-CoV-2 vaccines right now appear to be very effective against the disease severity and deaths, no vaccine is perfect [15].

Furthermore, two studies in Scotland and England confirm high protection rates (80%– 91%) after the first dose of Pfizer or Oxford/AstraZeneca vaccines [3,4]. Previous studies have found that a single dose of the Sputnik V vaccine may be enough to elicit a strong antibody response against SARS-CoV-2 [16,17]. However, In the United States, the U.S. Center for Disease Control and Prevention (CDC) reported a total of 10,262 SARS-CoV-2 vaccine breakthrough infections from 46 U.S. states and territories as of April 30, 2021[18]. A recent study reported that the effectiveness of the vaccines against infections had decreased from 91.7% to 79.8% between 3 May and 25 July in New York [19]. Additionally, a study published by National Healthcare Safety Network (NHSN) found that the two mRNA (nucleoside-modified) vaccines (Pfizer-BioNTech and Moderna) were effective by 74.7% in nursing home residents between March and May, however, the protection has declined to 53.1% between June and July[20].

Investigating the trend of SARS-CoV-2 infection severity, hospitalizations rate, and deaths among persons who got the first dose of the SARS-CoV-2 vaccine are urgently needed to support decision-making logistics such as cold chains, vaccination schedules, and follow-up.

## Material and methods

We collected SARS-CoV-2 infection and mortality rate data by the NCDC daily reports from 30 April 2021 until 15 July 2021. The study includes all patients who were admitted to hospitals with confirmed SARS-CoV-2 by repeat reverse-transcriptase–polymerase-chain-reaction assays (RT-PCR). The data were collected from the medical record of the patients by intensive care physicians and healthcare centers staff working in 34 healthcare centers and hospitals from different cities. Inclusion criteria were all vaccinated and unvaccinated cases admitted to the healthcare centers during this period. Concurrently, we tracked the number of people who received the first dose of the SARS-CoV-2 vaccine since 10 April 2021.

### Ethics statement

The study was approved by the Ethics Committee (National committee for Biosafety and Bioethics at Biotechnology Research Center, Tripoli - Libya, N°: BTRC-2021).

### First Dose of COVID-19 Vaccines

As the emergence of the new variant strains of SARS-CoV-2 B.1.617.2 (Delta) in the world in December 2020, they have led to a rapid increase in the circulation of the virus and a rising demands for vaccines.

UNICEF, through COVID-19 Vaccines Global Access (COVAX), supported the Libyan government by delivering 57,600 doses at 8 April 2021[21]. Also, as an intranational gift the Libya government received an around 200,000 doses of (Sputnik V) vaccines, and 150,000 doses of Sinovac vaccine on April 2021[12]. Thus, a countrywide mass vaccination campaign with the use of three different authorized vaccines (Oxford/AstraZeneca, Sinovac, Sputnik V) was conducted in Libya starting from 10 April, 2021[22]. Nearly 5000 doses were administered daily through 400 immunization centers in all cities of the country. So far, about 445,000 people have received the first dose of vaccines that provided by the national immunization program. Priority was given to frontline healthcare and hospitals workers, adults over 60, and patients with chronic underlying health conditions in all regions of the country.

Hance, 110,000 doses of Sinovac, 175,000 doses of Oxford/AstraZeneca, and 160,000 doses of Sputnik V were administered. The National Center for Disease Control (NCDC) has recommended that the Oxford/AstraZeneca vaccine to be used for people aged over 55 years old. The other vaccines (Sinovac and Sputnik V) to be used for people who is 16 years old or more.

Moreover, due to delays in providing vaccines throughout COVAX and countries who are producing the vaccines, the NCDC announced a deviation from the recommended protocol for the SARS-CoV-2 vaccines, prolonging the interval between doses from 2 to 4 months. This procedure had two advantages: The first, a longer gap between doses may improve the long-term immune response, as demonstrated by AstraZeneca’s vaccine[5]. Secondly, a larger number of elderly and people with chronic diseases will be vaccinate.

### Hospital Admissions after one dose of vaccines

As of 15 July 2021, approximately 445000 persons in Libya had been received the first dose of the SARS-CoV-2 vaccines.

According to the NCDC data from 30 April 2021 (W1) to 15 July 2021 (W22), the total number of patients who have a confirmed SARS-CoV-2 infection is 39996. 3179 out of the total (7.9%) were admitted to healthcare centers, and among those 226 patients (7.10%) died during this period.

At the same time, A total of 43 SARS-CoV-2 vaccine breakthrough infections (i.e. One dose obtained from the vaccine) were reported from 18 healthcare centers and hospitals as of 30 April 2021. Among these cases, 8 patients died. Patients in healthcare seem to be moving towards an average age of around 55-87 years old (Table 2). Notably, 23 patients who were admitted to healthcare centers are vaccinated with the CoronaVac (Sinovac) vaccine. 14 patients received the Oxford/AstraZeneca. While only two patients had the Sputnik V vaccine.

**Table 2.** Numbers of patients who were admitted to healthcare centers or hospitals after received one dose of vaccine in Libya. N: Number DM: Diabetic HT: Hypertension CoronaVac: Sinovac Astra: Oxford/AstraZeneca SputV: Sputnik v (-) = Data not available.

| **N** | **Healthcare center** | **N of patient (Type of vaccines)** | **N of died (Type of vaccine)** |
| --- | --- | --- | --- |
| **1** | Derna | 2 (CoronaVAC)<br>1 (Astra) | 1 (Astra) |
| **2** | Marj | 2 (CoronaVac) | 0 |
| **3** | Bayda | 1 (CoronaVac) | 0 |
| **4** | Sabha | 1 (Astra) | 1 (Astra) |
| **5** | Tripoli1 | 1 (Astra)<br>1 (CoronaVac)<br>1 (Sput V) | 1 (CoronaVac) |
| **6** | Tripoli 2 | - | - |
| **7** | Zliten | 3 (Astra) | 0 |
| **8** | Zawiya | - | - |
| **9** | Misrata | - | 0 |
| **10** | Zuwara | 2 (CoronaVac)<br>1 (Astra) | 1 (CoronaVac) |
| **11** | Tegy | 1 (Astra) | 0 |
| **12** | Tobruq | 2 (CoronaVac) | 1 (Corona Vac) |
| **13** | Ghadames | 2 (CoronaVac)<br>1 (Astra) | 0 |
| **14** | Benghazi | 2 (Astra) | 0 |
| **15** | Shakshok | 2 (CoronaVac) | 1 (CoronaVac) |
| **16** | Msallata | 3 (CoronaVac) | 1 (CoronaVac) |
| **17** | Khoms1 | 1 (CoronaVac)<br>1 (Astra) | 0 |
| **18** | Khoms2 | 1 (Sput V)<br>3 (CoronaVac)<br>2 (Astra) | 1 (CoronaVac) |
|  |  | **Total N of patient = 43** | **Total N of patient Died = 8** |

Of 445000 people who had taken the first dose of the vaccines, the percentage of patients who had been admitted to the healthcare centers, and died is very low (0.009, 0.001%) respectively.

Since July, the NCDC has reported initial evidence that people being infected with the Delta variant. Where the numbers testing positive increased five-fold, from 1600 the previous week to 15000 cases. It recorded its highest number of cases on 12 July (W20). However, the number of patients’ deaths remains lower compared to W11 (17-26 patients per day) (Fig.1).

**Figure 1.**
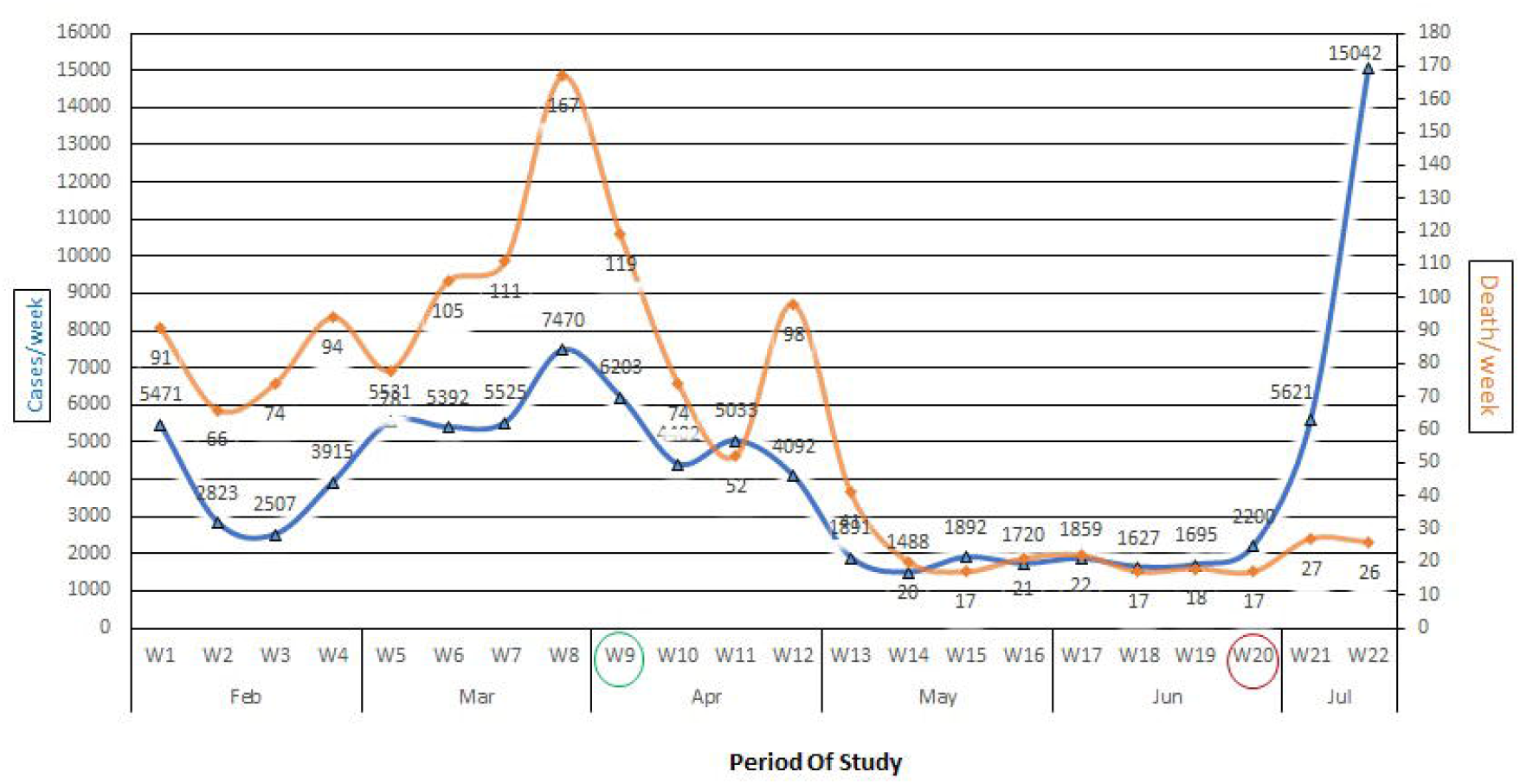
Number of patients confirmed SARS-COV-2 Infection from February to 15 July in Libya. Green ring: start to vaccines program Red ring: first cases with Delta variant.

In this period (W20), with delays in travel restrictions, there has been a significant increase in the number of people confirmed with SARS-CoV-2. Particularly in the western cities such as (Zawiya, Tripoli, Shakshok, Zliten, and Misrata), due to proximity to the Tunisian borders, which was recording the peak of infection rates in Africa and in the world with Delta variant (Fig 2).

**Figure 2.**
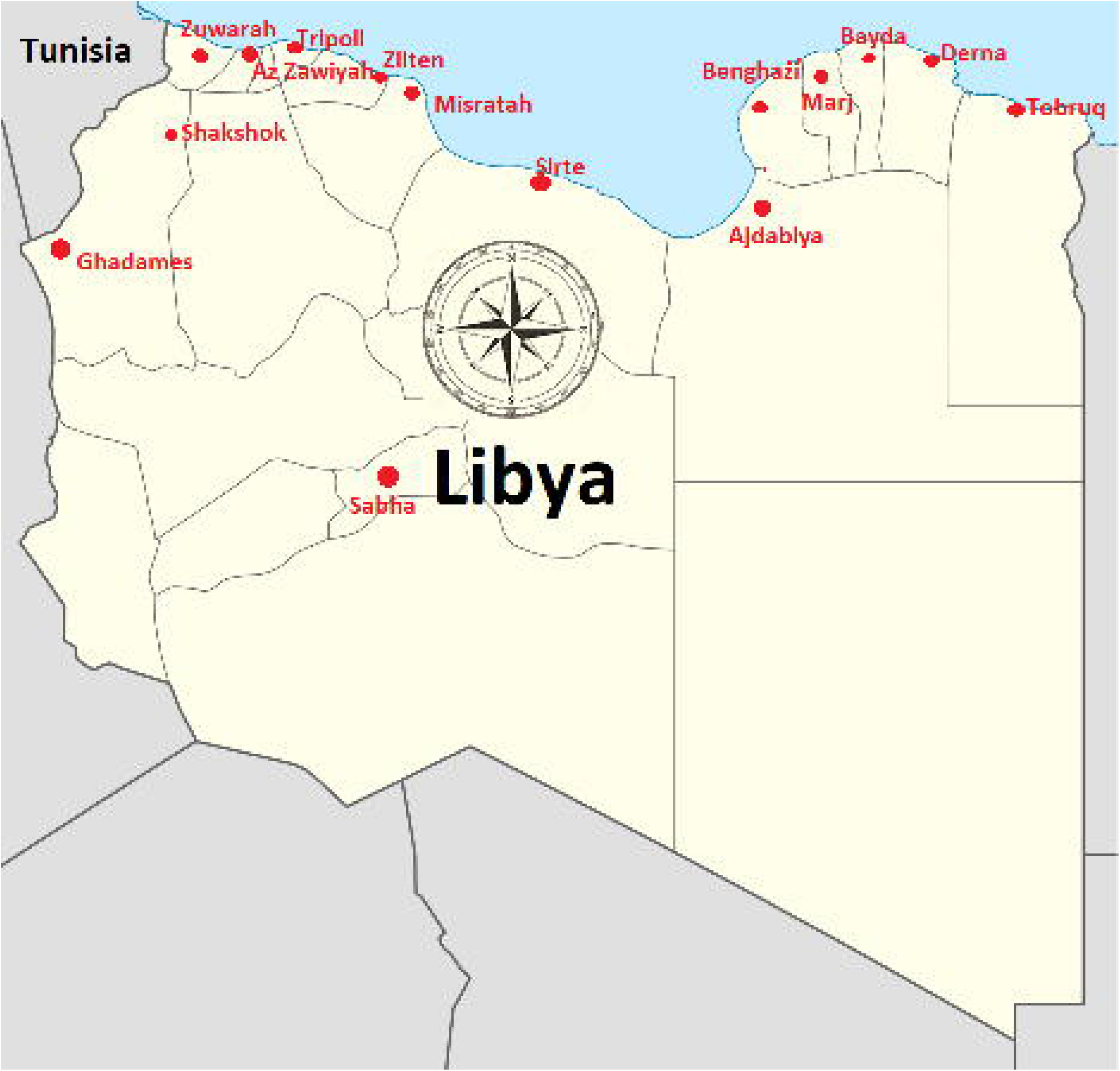
The geographic distribution of SARS-COV-2 infection in Libya.

### Limitations of study

The results in this study are subject to two limitations, the first one, that the number of reported SARS-CoV-2 vaccine breakthrough cases is substantially lower than that of all SARS-CoV-2 infections among one dose vaccinated persons. However, many people with vaccine breakthrough infections, especially those who are asymptomatic or who experience mild illness, might not seek testing. The second, SARS-CoV-2 sequence data are available for only a small proportion of the reported cases.

## Conclusion

In the present study, one dose of the three different types of vaccines had showed a decreasing in infection and mortality rate. The results were very encouraging. However, all these results were before the delta variant entered Libya. So as a recommendation, the department of health and NCDC must focus and make all possible efforts to get as many people vaccinated within a short period of time to prevent the development of new virus variants.

## Data Availability

All data were referred to in the manuscript

## Acknowledgements

The authors wish to thank all doctors in healthcare centers and hospitals who were involved in this study, and Hamza El-thelb for editing the text.

## Contributors

(I)Manuscript writing: All authors; (II) Final approval of manuscript: All authors.

